# Efficacy of commercial mouth-rinses on SARS-CoV-2 viral load in saliva: Randomized Control Trial in Singapore

**DOI:** 10.1101/2020.09.14.20186494

**Authors:** Chaminda J. Seneviratne, Preethi Balan, Karrie Ko Kwan Ki, Nadeeka S Udawatte, Deborah Lai, Dorothy Ng Hui Lin, Indumathi Venkatachalam, Jay Lim Kheng Sit, Ling Moi Lin, Lynette Oon, Goh Bee Tin, Jean Sim Xiang Ying

**Author notes:** Corresponding authors Associate Prof. Chaminda Jaya Seneviratne, Lead Principal Investigator, Singapore Oral Microbiomics Initiative, National Dental Research Institute Singapore (NDRIS), National Dental Centre Singapore, SingHealth Oral Health ACP, Duke NUS Medical School, Singapore, Dr. Jean Sim Xiang Ying, Consultant, Department of Infectious Diseases, Singapore General Hospital, SingHealth, Singapore. Equal contribution (Chaminda J. Seneviratne, Preethi Balan, Karrie Ko Kwan Ki, Jean Sim Xiang Ying).

## Abstract

The presence of high SARS-coronavirus 2 (SARS-CoV-2) titres in saliva may result in transmission of the virus and increase the risk of COVID-19 infection. This is particularly important as significant amounts of aerosols are generated during dental procedures, posing risk to dental care personnel and patients. Thus, reducing the titres of SARS-CoV-2 in the saliva of infected patients could be one of the key approaches to reduce the risk of COVID-19 transmission during dental procedures. In this randomised control trial, the efficacy of three commercial mouth-rinse viz. povidone-iodine (PI), chlorhexidine gluconate (CHX) and cetylpyridinium chloride (CPC), in reducing the salivary SARS-CoV-2 viral load in COVID-19 positive patients were compared with water. A total of 36 COVID-19 positive patients were recruited, of which 16 patients were randomly assigned to four groups— PI group (n=4), CHX group (n=6), CPC group (n=4) and water as control group (n=2). Saliva samples were collected from all patients at baseline and at 5 min, 3 h and 6 h post-application of mouth-rinses/water. The samples were subjected to SARS-CoV-2 RT-PCR analysis. The fold change of Ct values were significantly increased in CPC group at 5 minutes and 6 h time points (p<0.05), while it showed significant increase at 6 h time point for PI group (p<0.01). Considering Ct values as an indirect method of arbitrarily quantifying the viral load, it can be postulated that CPC mouth-rinse can decrease the salivary SARS-CoV-2 levels within 5 minutes of use, compared to water rinsing. The effect of decreasing salivary load with CPC and PI mouth-rinsing was observed to be sustained at 6 h time point. Within the limitation of the current study, it can be concluded that use of CPC and PI formulated commercial mouth-rinses, with its sustained effect on reducing salivary SARS-CoV-2 level, may be useful as a pre-procedural rinse to help reduce the transmission of COVID-19.

## Introduction

The Coronavirus Disease 2019 (COVID-19) pandemic caused by novel beta-coronavirus, has infected more than 24 million people with 841, 335 deaths globally (Worldometer 2020). At present, therapeutic strategies and preventive vaccines for SARS-CoV-2 have not been developed. Infection control strategies targeted at individuals as well as public health measures are key to curb the spread of SARS-CoV-2 infection.

Human-to-human transmission of SARS-CoV-2 have been reported to occur via droplets or contact transmission (Wu and others 2020). Hence in order to prevent cross-contamination, infection prevention practices such as hand hygiene, wearing mask and social distancing have been the mainstay of public infection control. A previous study has detected the presence of SARS-CoV-2 in the saliva of 91.7% COVID-19 patients, with a median viral load of 3.3 × 10^6^ copies/mL (To and others 2020). SARS-CoV-2 RNA in saliva from infected individuals have been found to be stable at 4°C, room temperature (∼19°C), and 30°C for prolonged periods (Ott and others 2020). Moreover, the detection rate of SARS-CoV-2 has been reported to be higher in saliva than nasopharyngeal swabs (Wyllie and others 2020). Thus, saliva can carry a risk of transmission of COVID-19, either via direct contact or indirect contact with contaminated objects Since the beginning of the pandemic, there has been a growing concern on the risk of SARS-CoV-2 transmission in dental practice (Meng and others 2020). Dental care professionals are exposed to aerosols from the oral cavity of patients, which can be a potential hazard to operator, dental auxiliary and other patients (Izzetti and others 2020). According to the World Economic Forum analysis, dental hygienists, dental assistants and dentists are among the occupations with highest COVID-19 risk (Lu 20 April 2020). Thus, reducing the salivary viral titers in COVID-19 patients could be one of the key approaches to prevent transmission of COVID-19, particularly in the dental settings.

The use of anti-septic mouth-rinsing has been suggested as a pre-procedural infection control measure by international as well as local health authorities. For instance, during the early stage of COVID-19 pandemic, Chinese health authorities recommended the use of Povidone-iodine (PI) and hydrogen peroxide based mouth-rinses as a pre-procedural preventive measure (Peng and others 2020), while National Dental Centre Singapore has advocated the use Cetylpyridinium chloride (CPC) mouth-rinse. However, the foregoing recommendations were not evidence-based as no clinical or *in-vitro* data is currently available on the effect of mouth-rinses on SARS-CoV-2. The first case report on efficacy of mouth rinses in reducing the SARS-CoV-2 viral load in the saliva was reported in Korea (Yoon and others 2020). Subsequently, Martínez Lamas et al., 2020 in a case-series of four patients suggested that PI mouth rinse could reduce the saliva viral load of SARS-CoV-2 in patients with higher viral loads (Martínez Lamas and others 2020). A recent *in-vitro* study has shown the efficacy virucidal activity of certain commercially available mouth rinses against SARS-CoV-2 (Meister and others 2020). Given that there are no clinical trials in the literature that have examined the efficacy of mouth-rinses to reduce the SARS-CoV-2 viral load in saliva, we evaluated the efficacy of three commercially available mouth-rinses, namely PI, CHX and CPC, on the salivary SARS-CoV-2 viral load in a cohort of COVID-19 positive patients in Singapore.

## Methods

### Patient cohort

A total of 36 laboratory-confirmed COVID-19 positive patients were recruited from Singapore General Hospital (SGH) from June 2020 to August 2020. History of allergy to PI, CPC or CHX and its relevant excipients, all forms of thyroid disease or current radioactive iodine treatment, lithium therapy, known pregnancy, and renal failure were considered as exclusion criteria. Ethics approval was obtained from SingHealth Centralized Institutional Review Board (CIRB Ref No: 2020/2537). The trial has been registered with ISRCTN (ISRCTN95933274). All patients provided informed consent upon recruitment in the study. The demographic characteristics of the subjects is depicted in Table 1.

**Table 1.** Demographic characteristics of subjects

|  | PI (n=4) | CHX (n=6) | CPC (n=4) | Water (n=2) |
| --- | --- | --- | --- | --- |
| Age (years) | 40.7 ± 11.5 | 43.6 ± 8.6 | 35.7 ± 8.5 | 36 ± 14.1 |
| Gender |  |  |  |  |
| *Male* | 4 (100%) | 6 (100%) | 4 (100%) | 1 (50.0%) |
| *Female* | 0 (0%) | 0 (0%) | 0 (0%) | 1 (50.0%) |
| Ethnicity |  |  |  |  |
| *Indian* | 3 (75.0%) | 4 (66.6%) | 3 (75.0%) | 2 (100%) |
| *Chinese* | 0 (0%) | 2 (33.3%) | 1 (25.0%) | 0 (0%) |
| *Others* | 1 (25.0%) | 0 (0%) | 0 (0%) | 0 (0%) |
| Comorbidities |  |  |  |  |
| *Present* | 1 (75.0%) | 1 (16.6 %) | 2 (50.0%) | 1 (50.0%) |
| *Absent* | 3 (25.0%) | 5 (83.3 %) | 2 (50.0%) | 1 (50.0%) |
| Time lapse between COVID-19 diagnosis and sample collection (days) | 2.2 ± 1.2 | 3.5 ± 0.8 | 2.5 ± 1.7 | 2 ± 0 |

### Sample collection

The enrolled patients were randomized using Robust Randomization App (RRApp) using block randomization technique (Tu and Benn 2017) and were allocated to four groups accordingly— PI, CHX, CPC and water control group. Prior to saliva collection, patients were asked to refrain from eating, drinking, or performing oral hygiene procedures for at least 30 min. Three milliliters of saliva was collected by passive drool technique from all the enrolled COVID-19 patients at four time points as described previously (Azzi and others 2020). Firstly, baseline saliva sample was collected prior to intervention of the mouth-rinse. Immediately after this, patients were requested to rinse their mouth with the allocated mouth-rinse for 30 seconds. Commercial mouth-rinses were prepared at the dilution and dosage recommended by respective manufacturers. In brief, PI group rinsed their mouths with 5 ml of PI mouthwash (commercially available as Betadine Gargle and Mouthwash 10 mg) diluted with 5 ml of water (0.5%) whereas the CHX group used 15 mL of undiluted CHX mouthwash (commercially available as Pearly White Chlor-Rinse, 0.2% w/v). The CPC group and water control group rinsed their mouths with 20 ml of 0.075% CPC (commercially available as Colgate Plax mouthwash) and 15 ml sterile water, respectively. Three milliliters of saliva were collected again from all subjects five minutes after the use of mouth-rinse. In order to evaluate the duration of the efficacy of mouth-rinses, salivary samples were collected at the 3 h and 6 h post-rinsing using the methodology described earlier.

### RT-PCR assay for SARS-CoV-2

Collected salivary samples of COVID-19 patients were immediately transported to the Molecular Laboratory, SGH. The SGH Molecular Laboratory performs routine diagnostic testing of COVID-19 using a validated SARS-CoV-2 reverse-transcription polymerase chain reaction (RT-PCR) assay. In brief, the in-house RT-PCR method used primers and probe from the protocol by Corman V et al, 2020 released by the World Health Organization (WHO) as well as Chu et al, 2020 protocol from the University of Hong Kong (Chu and others 2020; Corman and others 2020). This assay involves a single-step RT-PCR targeting the E gene of SARS-CoV-2. An internal spiked-in Bioline control (Bioline RNA Extraction Control 670 Kit #BIO-38040) or bovine-diarrhea disease virus (BVDV) is added to the assay prior to total nucleic acid extraction. These internal controls serve as extraction and amplification control. A run is considered valid if the positive control yields positive detection and negative controls remain negative

### Data analysis

From the total of 36 COVID-19 positive recruits, 19 patients had undetectable SARS-CoV-2 RNA in the saliva samples and were excluded from the study. In addition, one patient was excluded due to non-compliance with study protocol. Thus a total of 16 subjects comprising of PI group (n=4), CHX group (n=6), CPC group (n=4) and water as control group (n=2) were analysed in this study. A longitudinal comparison of the absolute CT values among all the time-point within each group was carried out using Analysis of Variance, followed by post-hoc tests to compare the differences between the groups. Relative fold change analysis was carried out after transforming the data on a scale of 1 to 5. Fold change was then estimated on the transformed data by calculating the ratio between Ct values at each time point versus the Ct value at baseline for each patient (Ct_timepoint_ / Ct_baseline_). The average fold changes values thus obtained at 5 min, 3 h and 6 h time points for each of the mouth rinse groups were compared with fold change value at corresponding time-points of water group using Independent t-test. The p-value threshold for significance was set at <0.05.

## Results

The Ct values detected in all 16 patients were within the range of 15.64 to 34.58, with a mean value of 27.73 + 4.77. The mean Ct values for the three mouth-rinses and water control group were depicted in Figure 1. Whilst the trend looks promising, no statistical differences were obtained in the Ct values with regards to any time points in all the groups.

**Figure 1.**
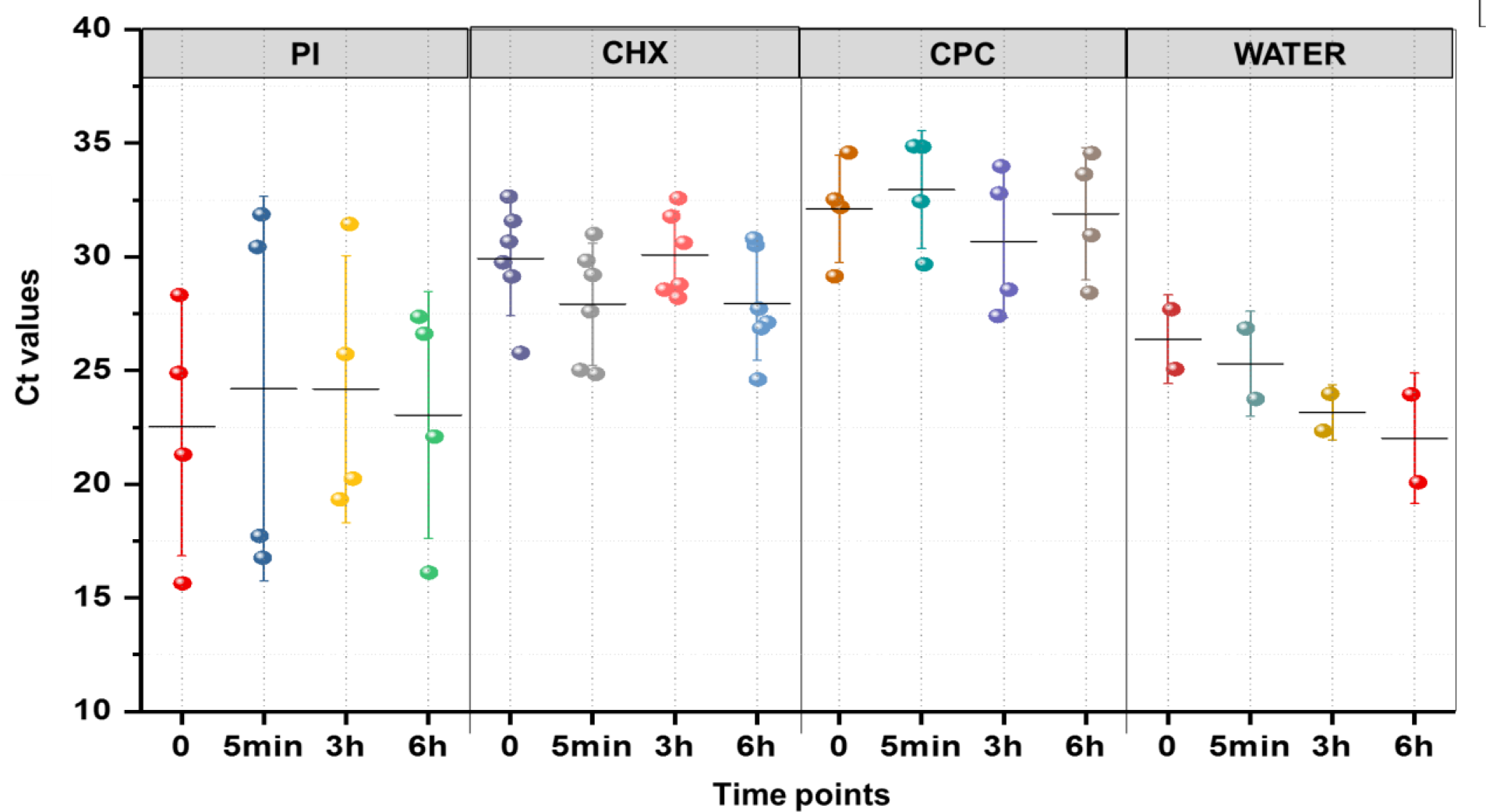
Cycle threshold (Ct) profile in saliva of COVID-19 positive patients treated with mouth rinses. Dots represents Ct value for each patient. Horizontal bar represents the mean value and vertical bar represents the 95 % confidence interval.

In order to compare the efficacy of the three mouth rinses, firstly the fold change of Ct value at each time point with respect to its baseline Ct value was estimated. Then the fold changes at 5 minutes, 3 h and 6 h in all the three mouth rinse groups were compared with the Ct value fold change at the respective time points of water group (Figure 2). A statistically significant increase in fold change of Ct value at 5 minutes and 6 h was observed post rinsing with CPC mouth rinse compared to the water group (p<0.05). Although the fold changes in Ct values were higher at 3 h in CPC group, no statistical significance was achieved (p=0.20). Similarly, the PI group also showed higher fold changes in Ct value post 5 minutes and 3 h of post-rinsing, compared to the water group. However, statistically significant increase in fold change was obtained only at 6 h post rinsing with PI in comparison with water (p<0.01). The CHX group demonstrated a varied effect among patients after 5 minutes rinsing and hence further studies with a larger sample size are required to determine its significance. Nevertheless, the trends at 3 h and 6 h post rinsing with CHX were consistent with other mouthwashes. Ct values are considered inversely related to viral load and therefore may serve as an indirect method of arbitrarily quantifying the viral load in the sample (Rao and others 2020). Hence, it can be postulated that CPC mouth-rinse decreased the salivary SARS-CoV-2 levels within 5 minutes of use, compared to water rinsing. The effect of decreasing salivary load with CPC and PI mouth-rinsing was observed to be sustained at 3 h and 6 h time points compared to the control group.

**Figure 2.**
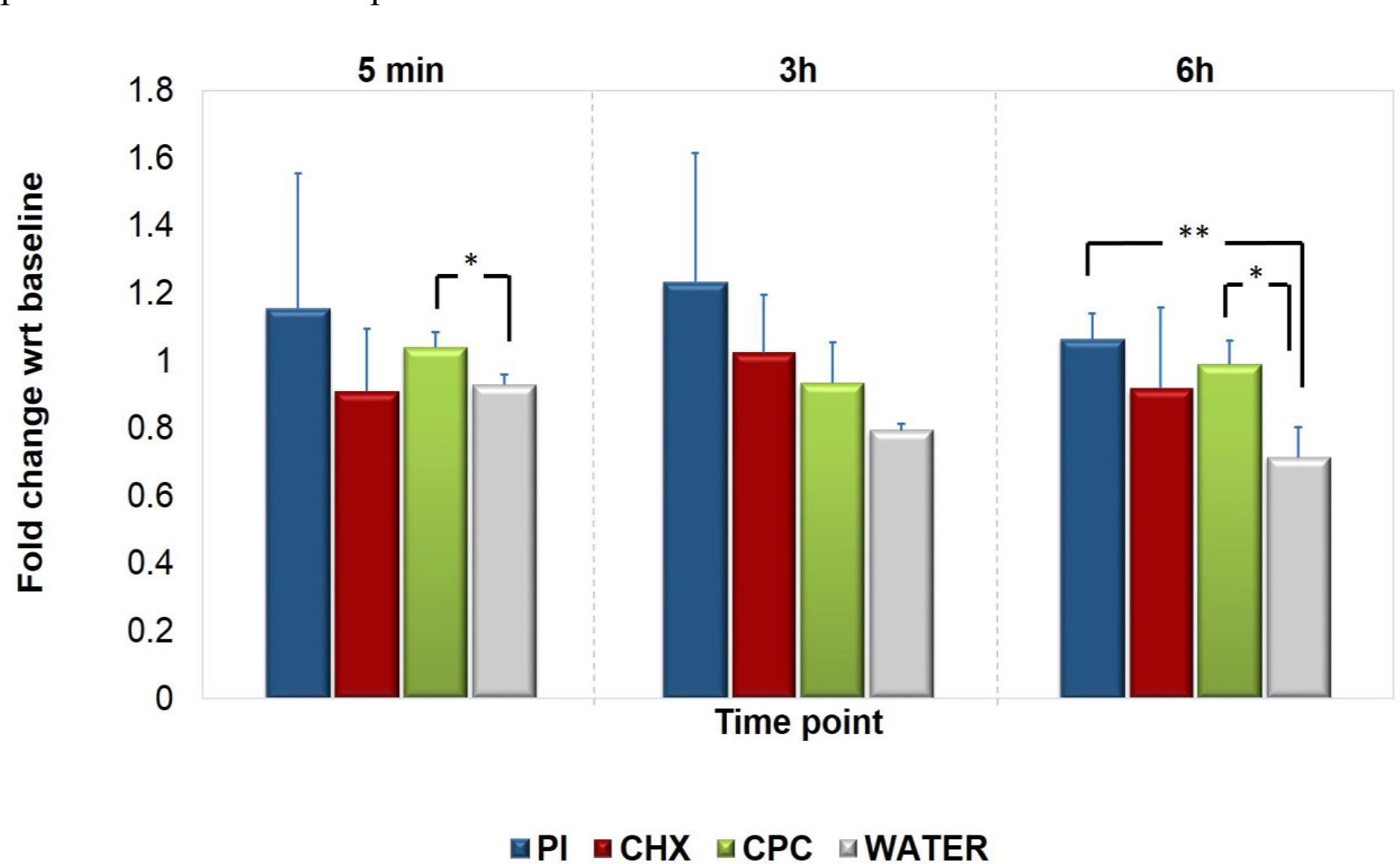
Relative fold change of Cycle threshold (Ct) values in mouth-rinse and water group. The fold change in Ct value at each time point is calculated with respect to its baseline Ct value. Each mouth-rinse group - Povidone-Iodine (PI), Chlorhexidine gluconate (CHX) and Cetylpyridinium chloride (CPC) were compared with water group using Independent t-test. ** represents P<0.01 and * represents P<0.05.

## Discussion

This is the first randomized clinical study to examine the efficacy of commercial mouth-rinses on SARS-CoV-2 in COVID-19 patients. In fact, only a single clinical trial is available in the literature that has evaluated the effect of essential oil containing mouth-rinses on Herpes simplex virus (Meiller and others 2005). In the aforementioned clinical trial, effectively zero recoverable virions were found at 30 s post rinse and the reduction in virus in saliva sustained significantly for 60 min, which was the last time point recorded. Hence, despite limitations, the present study will provide a novel insight on this important topic, particularly at the time of the current COVID-19 pandemic. It is noteworthy that anti-septic mouth-rinses should not be regarded as a treatment for SARS-CoV-2. Nevertheless, mouth-rinses could play a vital role in minimizing the spread of SARS-CoV-2 with its use as a pre-procedural strategy in dental clinics (Vergara-Buenaventura and Castro-Ruiz 2020). This is especially important as significant amounts of aerosols are generated in a relatively closed setting during dental treatments (Ge and others 2020). Health authorities such as, Centre for Disease Control and Prevention (CDC) and Australian Dental Association (ADA) have recommended the use of pre-procedural mouth-rinses for dental treatment, even without robust clinical evidence (ADA 2020; CDC 2020). Therefore, the present study provides the much-needed evidence on the efficacy of commercial mouth-rinses for salivary SARS-CoV-2 viral load in a group of COVID-19 positive patients in Singapore.

It was observed that CPC and PI mouth-rinses have sustained effect in reducing viral load in saliva compared to water control in our study. Previously, CPC has been shown to have anti-viral activity against influenza viruses in *in vitro* with an EC50 of 5–20 ug/ml and *in vivo*, through direct attack on the viral envelope (Popkin and others 2017). The virucidal spectrum of PI has been reported to be against both enveloped and non-enveloped viruses (Kawana and others 1997). An *in vitro* study has shown the antiviral effectiveness of a 0.12% CHX mouth-rinse on several viruses associated with the oral cavity (Bernstein and others 1990). Moreover, a recent *in vitro* study evaluated the virucidal activity of commercial mouth-rinses against SARS-CoV-2 (Meister and others 2020). This *in vitro* study found that three mouth-rinses containing different active components, namely dequalinium chloride/ benzalkonium chloride, polyvidone-iodine and essential oil could significantly reduce the SARS-CoV-2 viral load to undetectable level. However, chlorhexidine-based mouth-rinse failed to reduce the viral load significantly. O’Donnell et al. (2020) also suggested that chlorhexidine could only weakly inactivate coronavirus strains (O’Donnell and others 2020). In our study, we observed highly varied efficacy of CHX mouth-rinse on SARS-CoV-2 in saliva. Hence, we recommend further studies to establish the clinical efficacy of CHX in COVID-19 patients.

CPC is a quaternary ammonium compound which exerts its anti-viral effect through disruption of the viral lipid envelope through physicochemical interactions. Coronaviruses including SARS-CoV-2 are surrounded by lipid membrane or “envelop” (Alexander E Gorbalenya 2020). The spike glycoprotein responsible for the infection is inserted into this lipid envelop. Therefore, interruption of this lipid membrane by ethanol has been the key infection control strategy against SARS-CoV-2 transmission. As CPC and PI are able to destroy the lipid membrane of the SARS CoV-2, mouth-rinsing could be a safe, effective strategy to reduce the viral transmission through oral route. Interestingly, previous randomized clinical trial has shown that oral topical administration of ARMS-I, that contains CPC, to be effective in reducing severity and duration of upper respiratory tract infections in patients infected with viruses like influenza, coronavirus or rhinovirus. (Mukherjee and others 2017).

It has been suggested that the expression of angiotensin-converting enzyme II (ACE2), a receptor that play an important role in cellular entry of the SARS-CoV-2 virus, could be higher in minor salivary glands than that in lungs (Xu and others 2020b). Moreover, mucosa of oral cavity could also express the ACE2 receptors and was observed to be higher in tongue than other oral sites (Xu and others 2020a). Therefore, it is possible that oral epithelial cells may also contain the virus and the virus-infected oral epithelial cells may desquamate as a salivary component. Thus, it is postulated that virucidal activity of mouth-rinses may collectively reduce the viral infectivity in saliva as well as aerosolised viral particles from the oral route.

Although the evidence of the current study is encouraging, several limitations should be noted. The study was intended to recruit a larger cohort of COVID-19 patients based on the sample size calculation. However, recruitment was concluded after 36 patients due to drastic decrease in COVID-19 cases by late August, 2020 in Singapore. This limitation has also been reported in previous clinical studies looking into anti-viral effect of oral topical products (Mukherjee and others 2017). While the SARS-CoV-2 RT-PCR, employed in this study, was performed using a standardized protocol to enable viral load estimation, the assay was not designed to be a quantitative test. Moreover, the current testing method (RT-PCR) have the limitation of not being able to determine the viability of viruses. Given the difficulties in culturing SARS-CoV-2 virus from clinical specimens, using viral RNA load as a surrogate remains plausible at this moment (Joynt and Wu 2020). Therefore, more studies are warranted with a larger sample size and viral culture to arrive at a definitive conclusion on the efficacy of mouth-rinses on the SARS-CoV-2 viral load in saliva.

Nevertheless, within the limitations of the present study, it can be concluded that CPC and PI formulated commercial mouth-rinses may have a sustained effect on reducing the salivary SARS-CoV-2 level in COVID-19 patients. These mouth-rinses could be a useful pre-procedural infection control strategy in clinical dental settings, where aerosol generation is significant. Moreover, a substantial number of COVID-19 patients remain asymptomatic. In such situations, the routine use of antiseptic mouth-rinsing could be a cost effective approach in reducing viral outspread, with potentially low health risk. Considering that mouth-rinses are available over the counter, it holds potential as a strategy with high public health impact to minimize the transmission of SARS-CoV-2 through oral route.

## Data Availability

Available on request.

## Acknowledgements

This study was supported by the NMRC Centre Grant Seed Funding Program, National Dental Centre Singapore (NDCS) Research fund (133/20) and National Dental Research Institute Singapore (NDRIS) fund (11/FY2019/G1/02-A44) to CJS.

## Author contributions

CJS, PB, KKKK, JSXY contributed to conception, design, data acquisition and interpretation, drafted and critically revised the manuscript. NU, DL, DNHL, IV and GBT contributed to design and data acquisition, drafted and critically revised the manuscript. JLKS and LML, LO contributed to data acquisition and interpretation, drafted and critically revised the manuscript. All authors read and approved the final manuscript.

